# Effect of an Intensive Community-Based Next-Generation NeuroAnimation Therapy in Reducing Upper Extremity Impairment after Stroke: Small Retrospective Cohort Study

**DOI:** 10.64898/2026.06.26.26356720

**Authors:** Valerie A. Hill, Sarah Anderson, DJ Capetillo, Anna Pittman, Caroline Bourchard, Preston Nutwell

**Affiliations:** Division of Occupational Therapy, School of Health and Rehabilitation Sciences, College of Medicine, The Ohio State University, Columbus, OH; NeuroAnimation Brain Growth and Recovery Center, New Albany, OH

**Keywords:** Community-Based, High Intensity, Virtual Reality, NeuroAnimation Therapy, Upper Extremity Impairment, Stroke, Cohort Study

## Abstract

**Background:** Post-stroke motor impairment is the leading contributor to long-term disability. Despite evidence that high dose, high intensity (HDHI) and virtual reality (VR) interventions are effective in reducing post-stroke motor impairment, access to such interventions is limited, especially in community-based models. The purpose of this study was to explore the effect of one community-based HDHI VR intervention, Next-Generation NeuroAnimation Therapy (NG-NAT), on motor impairment for community-dwelling stroke survivors.

**Methods:** The study employed a retrospective pre-test post-test design of de-identified data sets of one cohort of stroke survivors who participated in an HDHI NG-NAT intervention at a community-based center from March to December 2025. The intervention consisted of three hours of daily therapy, five days a week, for three weeks. Two hours were allocated for NG-NAT gameplay, while one hour focused on non-VR activity. The NG-NAT was provided in a small studio with a large screen monitor and 12 motion caption cameras mapping client movements to play the game. The upper extremity Fugl Meyer Assessment was used to measure motor impairment at pre- and post-testing. Linear regressions were run to determine the relational strength between pre- and post-UEFMA scores. Wilcoxon Signed Rank Tests were run to calculate median differences in pre- and post-UEFMA scores and account for non-parametric data distributions at baseline and the small sample size. Effect size was explored using the Rank Biserial Correlation. Frequency of minimally clinically important differences (MCID), minimal detectable changes (MDC), recovery stage transition were calculated. Content analysis and co-review of documentation contextualized statistical findings.

**Results:** Nineteen participants completed three weeks of intensive NG-NAT. All experienced positive UEFMA score improvements from pre- to post-testing with a median difference of 8 points. Fifteen achieved MDC and MCID; one experienced a ceiling effect. Eight participants transitioned into better recovery stages. There was a highly significant, positive relationship with narrow confidence intervals and pre-score predicted post-score (e.g., those with mild/moderate impairment improved better than those with severe impairment).

**Conclusion:** This study provides evidence supporting the efficacy of NG-NAT as a community-based intervention to reduce motor impairment for individuals with stroke. Given its ability to deliver intense and engaging therapy, NG-NAT offers a promising adjunctive strategy to expand access for stroke survivors to improve clinically relevant health outcomes. These findings underscore the need for pragmatic trials evaluating effectiveness, implementation, and cost-effectiveness.

---

For over three decades, stroke has been a leading cause of long-term disability,^1,2^ specifically due to motor dysfunction.^3^ Despite evidence that high dose, high intensity (HDHI) training reduces motor impairment in stroke, implementation is rare in traditional rehabilitation models due to systematic healthcare barriers.^4^ Technological advancements, i.e., gaming, robotics, allow supplementation with traditional rehabilitation. *The purpose of this study was to retrospectively explore the effect of a HDHI intervention using an enhanced gaming system, Next-Generation NeuroAnimation Therapy (NG-NAT), on motor impairment with a small cohort of stroke survivors in a community-based setting*.

## Background

### Rehabilitation after Stroke

High intensity interventions effectively reduce motor impairment and improve activity engagement post-stroke.^5, 6^ Higher therapy dosage reduces rehospitalizations and improves activity engagement and motor function after stroke.^6, 7, 8^ However, traditional stroke rehabilitation—focused on facilitating functional independence, community mobility, and motor relearning—are provided at lower doses and intensities.^4^ Specifically, 24% of stroke survivors are discharged from acute care to inpatient rehabilitation, 27% to skilled nursing, 1% to long-term care, and 47% directly home.^9^ The frequency of home and outpatient therapy visits of stroke survivors on Medicare were analyzed in the STRONG study (n=510 patients across 28 medical systems).^10^ The median frequencies of therapy visits were exceptionally low at 0–1.5 and 6-8 sessions during the 3-12 months post-stroke, while 31% received no therapy.^10^

### Technological Intervention Approaches

Luckily, rehabilitation technologies can supplement therapeutic engagement and resultant functional gains by stroke survivors. In fact, varied therapeutic game-based technologies reduce motor impairment, e.g., robot-assisted gaming,^11^ non-immersive virtual reality (VR),^12^ and virtual exercise equipment.^13^ In a meta-analysis,^14^ 135 studies were synthesized comparing VR with alternative therapies, VR was more beneficial for UE function (moderate effect) and reducing activities limitations (small effect); and higher doses resulted in greater improvements.^14^ The authors deduced that gamification of therapy was the active ingredient that fostered enhanced engagement and positive outcomes for stroke survivors.

### Non-immersive NAT

NAT pairs motion capture with a non-immersive, VR game requiring intense cognitive-motor engagement of players. During gameplay, three-dimensional movements of the upper extremity (UE) control a virtual oceanic character (octopus) navigating underwater environments to complete goal-directed tasks (e.g., avoiding threats). Tasks were designed to elicit multi-planar UE movements across the active range of motion and were calibrated to progress difficulty based on performance (https://www.neuroanimation.com). Although not superior to time-matched occupational therapy intervention, NAT administered during outpatient rehabilitation facilitated motor improvement for clients with stroke (SMARTS2, Clinical Trial NCT02292251).^12^ Compared to traditional motor recovery, which targets cortical activation, MRI studies revealed glial growth of brain regions and improved cognitive efficiency after community-based NAT.^15^ The mechanism facilitating cognitive efficiency was neuronal growth in hippocampal regions— left subiculum, CA3 subregions.^15^ NAT has been feasibly implemented across care settings.^12, 15, 16^

#### Next-Generation NAT (NG-NAT)

NG-NAT—an enhanced version—offers refined motion capture fidelity and precision joint tracking to enable continuous fluid 3-dimensional movements (i.e., moving limbs up-down, flexion-extension of fingers). Addressing the gap in stroke rehabilitation and the need for HDHI rehabilitation business models,^4^ NG-NAT was established in a community-based model—Neuro Recovery Center (the Center)—beginning in 2025. *The purpose of this study was to explore the effect of NG-NAT on motor impairment for stroke survivors who participated in the Neuro-Recovery Bootcamp (NRB). We hypothesized that motor impairment would significantly improve from pre-to post-testing*.

## Methods

This cohort study employed a retrospective pre-test post-test design of de-identified data sets of clients with stroke who participated in NRB at an NAT center from March to December 2025.

### Research Ethics

This study was deemed exempt from ethics review by the corresponding author’s institutional review board on 1/25/2026. Specialists administered UE Fugl Meyer Assessment (UEFMA) from May until December 2025 on Day 1—pre-evaluation, and Day 15—last therapy session. Data was collected by the industry partner, de-identified, and shared with researchers. Data included participant UEFMA scores at pre- and post-testing, client goals, therapy notes, and relevant demographics, e.g., stroke diagnosis. Non-UEFMA data were used to contextualize statistical results.

### Outcome Measure

UEFMA was administered by NG-NAT specialists at the first and last NRB sessions. A psychometrically sound assessment, the UEFMA was used to quantify motor impairment based on stroke recovery motor patterns, coordination, and speed.^17^ With 33 items and 66 total points, movements were scored on a 3-point Likert scale, total (0), partial (1), and none (2). Motor impairment severity was categorized into three groups, severe, moderate, and mild.^18^ To reduce measurement bias, specialists used a rigorous, standardized UEFMA protocol. Initial inter- and intra-rater reliability was confirmed across specialists. All UEFMA sessions were video-recorded and reviewed for administration validity and inter-rater reliability by NG-NAT supervisors. Discrepancies were reconciled between the specialists, and further training provided.

### Intervention

#### NRB-NG-NAT

NRB-NG-NAT comprised of NG-NAT and goal-directed activity (i.e., activity). Active ingredients included (1) coaching by NG-NAT specialists, (2) two hours of NG-NAT in a distraction-free environment, (3) one hour of activity in therapy gym, and (4) real-time multi-sensory feedback with progressive cognitive-motor challenge. The goal of NRB-NG-NAT was to increase the amount and quality of UE movement in challenging yet rewarding environments.

##### Specialists and Coaching

Specialists, exercise physiologists, received standardized training prior to leading NRB-NG-NAT. Specialists designed intervention plans based on client-identified goals, NRB-NG-NAT protocol, and the just-right-challenge. Quality movement was encouraged.

##### Delivery Model and Environment

NRB-NG-NAT was delivered 3-hours x 5-days/week x 3-weeks. NG-NAT occurred in the first and third hours in a theatre (e.g., 10x10 ft^2^ room) equipped with a large screen display (e.g., 10’ x 8’) and 12 motion-capture cameras around the room. Non-NAT Activity occurred in therapeutic spaces in the second hour.

##### Feedback and Challenge Progression

During NG-NAT, clients navigated characters in underwater games by performing complex motor movements. Visual and auditory feedback was provided throughout gameplay by the game and specialists. Specialists controlled the calibration and difficulty levels and adjusted targets in real time, increasing difficulty levels as movement patterns improved, promoting quality movement rather than compensatory strategies when using affected UE.

During Activity, clients performed tasks to facilitate strength, coordination, and function needed to achieve their goals. Activities focused on upper and lower extremity strengthening and stretching exercises; dual-task training; advanced coordination; walking; stepping and stairs; basic and instrumental activities of daily living (ADLs); active hobbies (e.g., fishing, hiking); and creative hobbies (e.g., painting).

### Data Analysis Plan

Descriptive statistics were run for UEFMA measures of central tendency, range, and standard deviations. Data distributions were tested with Shapiro Wilk. Linear regressions were run to determine the relational strength between pre- and post-UEFMA scores. Wilcoxon Signed Rank Test was run to calculate median differences in pre- and post-UEFMA scores and account for non-parametric data distributions at baseline and the small sample size. Effect size was explored using the Rank Biserial Correlation. Analyses were set with alpha=0.05 and one-sided hypothesizing that scores would improve. Frequency of minimally clinically important differences (MCID) and minimal detectable change (MDC) of greater than or equal to 5.25 points were calculated.^19, 20^ Frequency of recovery stage transition was calculated based on stroke motor impairment categories of severe (0-28), moderate (29-42), and mild (43-66).^18^ Content analysis of treatment documentation and co-review of notes with specialists were conducted to contextualize statistical findings.

### Procedures

#### Data collection

Participants were assessed on the first and last days of NRB. Data was provided to the research team (February 2026), checked and cleaned (March), analyzed (April), and interpretation (May). Participant data sets were chosen for all stroke survivors who had completed the NRB prior to data retrieval.

Electronic health record (EHR) data was extracted by NRC staff into Microsoft Excel. The data was de-identified and transmitted to researchers via a password-protected email and saved to a secure password-protected folder in the University Medical Center OneDrive. Data was cleaned in Microsoft Excel and analyzed in Jamovi Cloud, an open access statistical software. Charts and figures were run to visualize data.

Source data verification was used to validate accuracy and completeness of raw data. EHR data and research dataset were double-verified by two research staff and two NRB-NG-NAT supervisors. Data differences were reconciled for three datasets that hadn’t been double checked for reliability.

## Results

Nineteen UEFMA data sets were collected; there was no missing data. NRG-NG-NAT occurred between April and December 2025 and participants completed all sessions. There was a wide age range (22-65 years) and most participants had an ischemic stroke more than six months prior to enrollment. All participants completed three weeks of intensive NG-NAT. None lived within driving range of the center and lodged at a local hotel. All participants paid out of pocket for services (approximately $8,000) and lodging, although discounts were available. Fifteen participants achieved MDC and MCID; one experienced a ceiling effect (pre-UEFMA: 65/66; post-UEFMA: 66/66; 1 point improvement). All experienced positive UEFMA score improvements from pre-to post-testing (M=8; range: 1-15). Motor impairment at baseline ranged from severe (12), moderate (4), and mild (3). Participants transitioned from severe to moderate (7) and moderate to mild (1) impairment levels (Table 1).

Participants showed an increase in scores from pre-test (M=26.9, Mdn=26, SD=16.357) to post-testing (M=34, Mdn=34, SD=15.611). A Shapiro-Wilk test determined the data were normally distributed for pre-testing (W=0.946, p=0.341) and post-testing (W=0.937, p=0.232) (Table 2, Figure 1).

A Pearson product-moment correlation matrix determined a highly significant, positive relationship (r=0.979); a strong linear slope with narrow confidence intervals (CI: 1.00, 0.952) (Table 3, Figure 2). A statistically significant linear regression found that for each point increase in pre-scores, the predicted post-scores increased by approximately 0.93 points (t= 19.701, p<0.0001) (Table 4).

A Wilcoxon signed-rank test determined that the median score significantly increased after the intervention (Mdn=8, W=0.00, p<0.0001, r=1.000) (Table 5).

## Discussion

This study was the first to retrospectively explore the effect of NG-NAT on motor impairment with a small cohort of stroke survivors (n=19) in a community-based setting. Despite the small sample size, statistical results indicated a large, significant treatment effect experienced by stroke survivors who participated in the NRB-NG-NAT. All participants experienced increased UEFMA scores; many achieving clinically meaningful thresholds (n=15/18; +5.25 points). Statistical analyses reinforced that motor recovery improved due to treatment, not by random chance. Although findings are not generalizable to the greater stroke population, this study adds preliminary evidence of efficacy of community-based HDHI gaming interventions for motor recovery. Notably, the intervention was implemented in a community-based business setting, uncommon in stroke rehabilitation. This study provides a model for reporting clinically meaningful, real-world outcomes in non-traditional rehabilitation settings.

### Clinical Relevance

The goal of NRB-NG-NAT—to increase the amount and quality of UE practice through meaningful and rewarding engagement—was highly successful for most participants. Most clients in this data set experienced real and meaningful reductions in motor impairment, making this clinically relevant. These findings support the fact that the statistical results are reliable and not due to measurement error.

Interestingly, motor recovery occurred consistently with Brunnstrom’s stages of recovery^21^ and Hijikata, et. al.’s findings of a Rasch analysis of UEFMA item difficulty. Proximal—shoulder and elbow—movements were easier for stroke survivors to perform, and forearm and wrist movements were more difficult.^22^ In fact, stroke survivors with moderate-severe impairment had the greatest reduction in impairment and transitioned from severe impairment to moderate impairment,^22^ as observed with case C38. This trend is visibly notable in Table 6, a matrix of UEFMA change scores for a sample of six cases.

### Case Contextualization

Six cases were selected to contextualize statistical and clinically relevant findings. Demographic information included average age 52 years old (range: 22-65), female (4), white (5), ischemic stroke (5), stroke onset <12 months (4), positive UEFMA change (range: 5-15 points), and achieved MDC and MCID (n=5/6 cases) (Table 6). All individuals completed 3 weeks of intensive NRB-NG-NAT. No participants lived within driving range of the center and lodged at a local hotel. All participants paid out of pocket for services (approximately $8,000) and lodging, although discounts were available.

Across client data sets, impairment was reduced across UE regions (shoulder, forearm, wrist & hand) and quality of motor control was improved (tremors, dysmetria, speed). Consistent with motor recovery stages,^21^ clients with lower pre-scores (e.g., 6/66) were unable to get into the test starting positions and scored 0, and at post-testing demonstrated ability to achieve starting positions and partial movement primarily for shoulder internal/external rotation and forearm supination/pronation. The client with a higher pre-score (e.g., 50), was able to get into starting positions, but unable to achieve full movement pattern. At post-testing, they were able to achieve the full movement pattern and there were improvements in coordination (i.e., tremor). There appears to be a trend that as motor impairment decreases, motor coordination improves, as we would expect in stroke recovery (Table 6).

Of the three cases who did not achieve MDC or MCID (5.25-point change), all had severe motor impairment. The client with the lowest pre-score (A27 [6 points]) was unable to achieve any starting positions at baseline. At post-testing they gained full movement for shoulder retraction (2 points), wrist flexion/extension (2 points) elbow flexion (1 point). A27’s goals were to improve independence in basic activities of daily living and walking endurance without an ankle foot orthosis (AFO), to grasp objects, and bend her knee. Goal-directed activities included task-specific training for ADLs, goal-directed strengthening for wrist extension, shoulder flexion/abduction, finger flexion, knee flexion/extension, walking in harness, and walking to metronome.

The case with the largest increase in UEFMA score (C38 [15 points]), had a lower baseline score (20 points)—indicating severe motor impairment. At baseline, C38 was able to get into one starting position (1 point), and unable for 9 positions (0 points). At post-testing, the client was able to get into starting positions, specifically for shoulder abduction/adduction, internal/external rotation, and flexion; forearm supination and pronation; and mass hand flexion. C38 also had improved motor coordination evidenced by a reduction in tremors and dysmetria. Significantly, this case transitioned into lower motor impairment severity (severe to moderate), primarily by being able to move their UE into starting positions and completing partial movement. C38’s goals included improving mobility and hand function for greater participation in active hobbies (e.g., golfing, fishing, pickleball) and improving slow and slurred speech. Goal-directed activities included playing adapted pickleball with both hands gripping the racket; stair climbing; navigating obstacle courses; dual-task training (describing and performing activities simultaneously); lower extremity (LE) strengthening and stretching exercises; and mindfulness techniques.

The case with partial baseline movements (E47 [baseline score: 32; post-testing score: 39; 7-point increase) was unable to get into starting positions at baseline and achieved partial movement for supination/pronation and wrist circumduction. They improved from partial to full movement for shoulder abduction at 90’ and external rotation, and wrist stability at post-testing. No changes were noted in the elbow or hand. E47’s goals were to increase left UE, wrist, and LE movement; manage foot drop and spasticity; and increase functional mobility independence (i.e., reduce reliance on adaptive devices). Goal-directed activities included kicking soccer balls in gravity-assist harness; hurdles and stair climbing without AFO; sequenced walking; standing balance; cognitive dual-task training; closed-chain UE wall activities; supported out-of-synergy movements (i.e., elbow and wrist extension); and grasp (gross and pincer) and finger dexterity activities.

The case with the highest baseline score (50 points) with significant improvement (10-points) gained full wrist and hand movements, and flexibility and strength to place hand on lumbar spine. C16’s goals were to increase use of left UE; improve walking and dancing; and return to work as an elementary school teacher. Goal-directed activities included step-by-step dance movement; hitting moving targets on screen; walking to metronome while speaking; cognitive dual-task training; functional exercises (e.g., kettle bell walk, stair step-ups, squats on balance cushion); and targeted strengthening exercises (e.g., shoulder raises, clamshells, hip abduction, grasp).

As noted in Table 1, goal-directed activities matched the participants’ goals and impairment levels. With the highly engaging HDHI NRB-NG-NAT and complimentary goal-directed activities, participants collectively progressed through the stages of motor recovery. It is anticipated that comparably resourced stroke survivors with similar function and goals would have a similar outcome.

### Implications

Based on these results— (1) highly statistically significant, (2) achievement of MDC and MCID, and (3) the promise of offering HDHI VR intervention (e.g., NG-NAT) in a successful community-based business model—hybrid effectiveness-implementation studies are the next step for clinical translation. With over 119 VR effectiveness studies, future studies need to analyze the usability and validity of VR interventions, while assessing effectiveness in real world settings.^14^ Dose and temporal optimization of HDHI interventions need to be explored for greater accessibility of precision stroke rehabilitation, particularly in non-traditional settings. For a comprehensive understanding of intervention effects, studies must analyze impairment level outcomes (e.g., cognition); brain function (e.g., hippocampal growth, neuroplasticity); activity and participation level outcomes (e.g., functional cognition, occupational performance, community reintegration), and factors impacting survivors access to evidence-based HDHI interventions (i.e., social determinants).

### Limitations

There was a risk of measurement bias for UEFMA administration. NAT specialists administered the UEFMA based on an evidence-based standardization protocol. All UEFMA were video recorded and double cross-checked for inter-rater reliability, substantially reducing potential bias. There was a risk of information bias with the exchange of data sets. To reduce bias, rigorous raw data checks and reconciliation methods were conducted with the research team and NAT director. There were inconsistencies in three data points, all of which were explained by a lag in inter-rater reliability checking and resolved at the raw data check.

### Conclusion

HDHI VR neurorehabilitation interventions are effective for individuals with post-stroke motor impairment. A new community-based delivery model—NG-NAT—shows promise for efficacious HDHI interventions with clinically relevant and highly significant effect on motor impairment reduction as evidence with a small cohort of stroke survivors. Hybrid effectiveness and implementation studies are warranted to explore pragmatic delivery approaches with rigorous methodologies.

## Supporting information

STROBE Checklist

## Data Availability

All data produced in the present work are contained in the manuscript

## Acknowledgements

Thank you to Omar Ahmad, PhD, Chief Executive Officer, Gaurav Gupta, Chief Operations Officer, and the NeuroAnimation Therapy Specialists of NeuroAnimation, Inc., for their collaborative partnership. Dr. Ahmad refined and implemented NG-NAT in the community-based business model and Gaurav Gupta led the research taskforce and collaboration between the research and center teams. NeuroAnimation Therapy Specialists allowed researchers to observe and discuss the intervention.

