## Supplemental Figures and Tables for "Effect of an Intensive Community-Based Next-Generation NeuroAnimation Therapy in Reducing Upper Extremity Impairment after Stroke: Small Retrospective Cohort Study"

**Tables and Figures**

Table 1. UEFMA Datasets (n=19)

| ID | Pre- UEFMA | Post UEFMA | Change Pre-Post | Achieved MDC & MCID |
| --- | --- | --- | --- | --- |
| 1 | 3 | 15 | 12 | Y |
| 2 | 6 | 14 | 8 | Y |
| 3 | 6 | 11 | 5 | N |
| 4 | 13 | 17 | 4 | N |
| 5 | 16 | 29 | 13 | Y |
| 6 | 19 | 22 | 3 | N |
| 7 | 20 | 34 | 14 | Y |
| 8 | 22 | 32 | 10 | Y |
| 9 | 24 | 34 | 10 | Y |
| 10 | 26 | 35 | 9 | Y |
| 11 | 27 | 33 | 6 | Y |
| 12 | 28 | 36 | 8 | Y |
| 13 | 29 | 37 | 8 | Y |
| 14 | 31 | 39 | 8 | Y |
| 15 | 34 | 40 | 6 | Y |
| 16 | 38 | 48 | 10 | Y |
| 17 | 50 | 60 | 10 | Y |
| 18 | 54 | 60 | 6 | Y |
| 19 | 65 | 66 | 1 | _*_ |
| _*Removed due to ceiling effect_ | | | | |

| Table 2. Descriptive Statistics  **Descriptives** | | | |
| --- | --- | --- | --- |
|  | Pre-UEFMA | Post-UEFMA | Change Pre-Post |
| N | 19 | 19 | 19 |
| Missing | 0 | 0 | 0 |
| Mean | 26.895 | 34.842 | 7.947 |
| Median | 26.000 | 34.000 | 8.000 |
| Mode | 6.000 | 34.000ᵃ | 8.000ᵃ |
| Standard deviation | 16.357 | 15.611 | 3.374 |
| Minimum | 3.000 | 11.000 | 1.000 |
| Maximum | 65.000 | 66.000 | 14.000 |
| Shapiro-Wilk W | 0.946 | 0.937 | 0.978 |
| Shapiro-Wilk p | .3406 | .2324 | .9194 |
| ᵃ More than one mode exists, only the first is reported | | | |

Figure 1. Histograms of UEFMA Scores

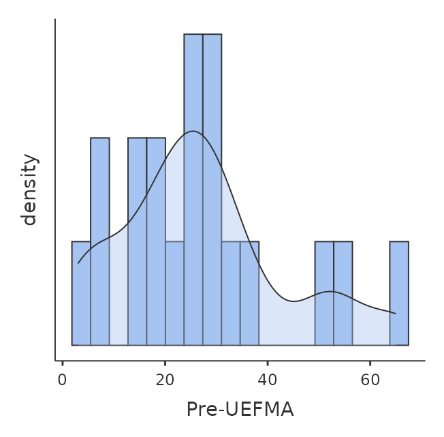

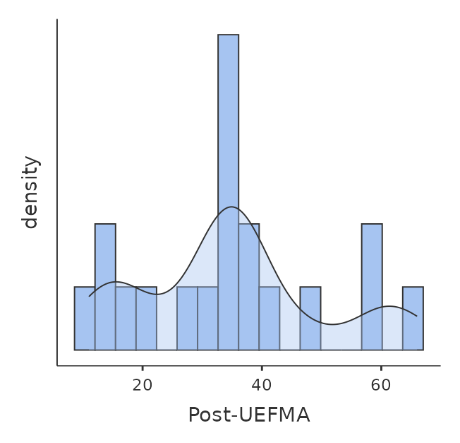

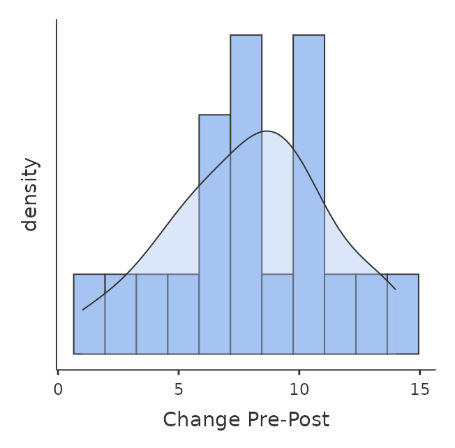

| Table 3. Correlation Matrix of Pre- and Post- UEFMA | | | |
| --- | --- | --- | --- |
|  |  | Pre-UEFMA | Post-UEFMA |
| Pre-UEFMA | Pearson's r | — |  |
|  | Df | — |  |
|  | p-value | — |  |
|  | 95% CI Upper | — |  |
|  | 95% CI Lower | — |  |
|  | N | — |  |
| Post-UEFMA | Pearson's r | 0.979 | — |
|  | df | 17 | — |
|  | p-value | <.0001*** | — |
|  | 95% CI Upper | 1.000 | — |
|  | 95% CI Lower | 0.952 | — |
|  | N | 19 | — |
| Note. Hₐ is positive correlation | | | |
| Note. * p < .05, ** p < .01, *** p < .001, one-tailed | | | |

Figure 2. Scatterplot of Pre- and Post-UEFMA

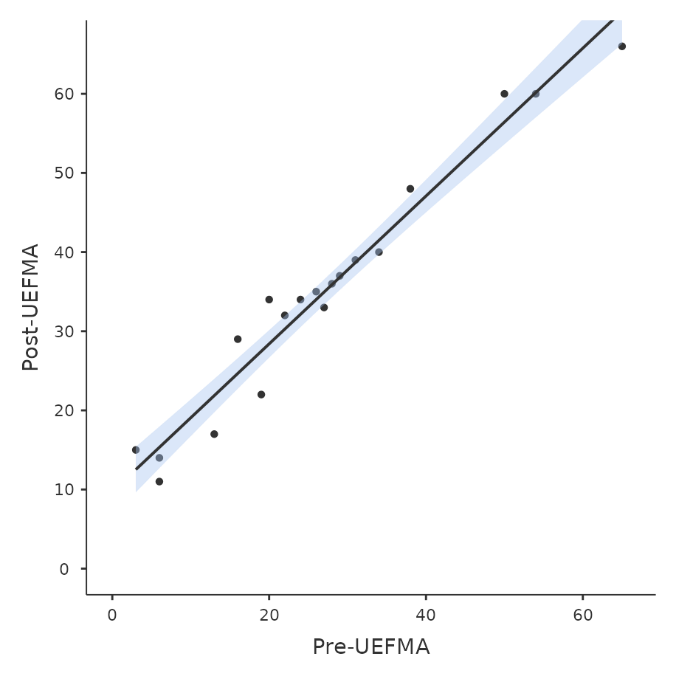

Table 4. Linear Regression of Pre-UEFMA (covariate) on Post-UEFMA (dependent variable)

| Model Fit Measures | | | | | | |
| --- | --- | --- | --- | --- | --- | --- |
| Model | | R | | R² | | |
| 1 | | 0.979 | | 0.958 | | |
| Note. Models estimated using sample size of N=19 | | | | | | |
| Model Coefficients - Post-UEFMA | | | | | | |
| Predictor | Estimate | | SE | | T | p |
| Intercept | 9.718 | | 1.482 | | 6.558 | <.0001 |
| Pre-UEFMA | 0.934 | | 0.047 | | 19.701 | <.0001*** |

Note. * p < .05, ** p < .01, *** p < .001, one-tailed

| Table 5. Wilcoxon Signed-Rank Test: UEFMA Median Differences from Pre- to Post-Testing | | | | | | | | | | | | |
| --- | --- | --- | --- | --- | --- | --- | --- | --- | --- | --- | --- | --- |
|  | | | | | | | 95% Confidence Interval | |  | | 95% Confidence Interval | |
|  |  |  | Statistic | p | Mean difference | SE difference | Lower | Upper |  | Effect Size | Lower | Upper |
| Pre | Post | Wilcoxon W | 0.000 | <.0001 | -8.000 | 0.774 | -Inf | -6.500 | Rank biserial correlation | -1.000 |  |  |
| Note. Hₐ μ_Measure 1 - Measure 2_ < 0 | | | | | | | | | | | | |

Table 6. UEFMA Scores and Subcategory Score Changes for Cases.

| **UEFMA** | **Case Examples** | | | | | |
| --- | --- | --- | --- | --- | --- | --- |
|  | **A27** | **B48** | **C38** | **D32** | **E47** | **F16** |
| Pre-Score | 6 | 6 | 20 | 25 | 32 | 50 |
| Post-Score | 11 | 14 | 35 | 32 | 39 | 60 |
| Change Score | 5 | 8 | 15 | 7 | 7 | 10 |
| MDC, MCID Achieved | N | Y | Y | Y | Y | Y |
| Subcategory Score Change (listed in testing order- Brannstrom recovery stages) | | | | | | |
| Shoulder Retraction | 0-2 | 1-2 |  |  |  | 1-2 |
| Elbow Flexion | 0-1 | 0-2 |  |  |  |  |
| Wrist Dorsiflexion @90*? | 0-1 |  |  |  |  | 1-2 |
| Wrist Volarflexion @90*? | 0-1 |  |  |  |  | 1-2 |
| Shoulder Elevation |  | 1-2 |  |  |  |  |
| Shoulder Abduction to 90* |  | 0-1 | 0-1 | 0-1 | 1-2 |  |
| Shoulder Adduction & IR |  | 0-1 | 0-2 |  |  |  |
| Mass Hand Flexion |  | 0-2 | 0-2 |  |  |  |
| Forearm Pronation/Supination (90*) |  |  | 0-1 | 1-2 | 0-1 |  |
| Forearm Pronation/Supination (30*) |  |  | 0-1 |  | 0-1 |  |
| Hand To Lumbar Spine |  |  | 0-1 |  |  | 1-2 |
| Thumb Adduction |  |  | 0-1 |  |  |  |
| Tremor |  |  | 1-2 |  |  | 1-2 |
| Dysmetria |  |  | 0-2 |  |  |  |
| Shoulder Flexion |  |  | 0-2 | 0-1 |  |  |
| Wrist Stability @15’ Ext Elbow @90* |  |  |  | 0-1 | 1-2 |  |
| Wrist Stability @15’ Ext Elbow @0* |  |  |  | 0-1 | 1-2 |  |
| Repeated Elbow Flexion/Extension @90* |  |  |  | 0-1 |  |  |
| Repeated Elbow Flexion/Extension @0* |  |  |  | 0-1 |  |  |
| Shoulder External Rotation |  |  |  |  | 1-2 |  |
| Wrist Circumduction |  |  |  |  | 0-1 |  |
| Wrist Dorsiflexion @0* |  |  |  |  |  | 1-2 |
| Wrist Volarflexion @0* |  |  |  |  |  | 1-2 |
| Hook Grasp |  |  |  |  |  | 1-2 |
| Spherical Grasp |  |  |  |  |  | 1-2 |
| Pincer Grasp |  |  |  |  |  | 1-2 |
| Shoulder Abduction |  |  | 1-2 |  |  |  |
| *Blank cells indicate no change. | | | | | | |

The jamovi project (2025). *jamovi*. (Version 2.7) [Computer Software]. Retrieved from [https://www.jamovi.org](https://www.jamovi.org/).
